# Food insecurity prevalence among tertiary students in Ghana

**DOI:** 10.1101/2024.02.27.24303448

**Authors:** Evelyn Agbetsise, Crystal Bubune Letsa, Charlotte Abra Esime Ofori, Geoffrey Adebayo Asalu

## Abstract

**Background:** Food insecurity (FI) is a collective term for inadequate access to available, affordable, and safe nutritious food. FI could affect students’ health and academic performance adversely. Currently, there is a paucity of research on food insecurity among tertiary students in Ghana. This study determined the prevalence of food insecurity among tertiary students in an Eastern city of Ghana.

**Materials and methods:** A descriptive cross-sectional study design was employed. A multistage sampling technique was used to select 411 respondents from four tertiary institutions. The United States Department of Agriculture Adult Household Food Security Survey (HFSS) Module tool was used to collect information on students’ food insecurity status. Data were entered with Epi-data version 3.0 software and then exported into Stata/MP version 16.0 for analysis. For all statistical tests, a p-value < 0.05 was considered significant.

**Results:** The prevalence of food insecurity was 61.0 %. Of these, 40.4 % had very low food security and 20.2% had low food security. Multiple logistic regression showed that the odds of food insecurity were significantly higher for students who did not have family support [AOR=2.56 (95% CI:1.17 - 5.63), p= 0.019] as compared to those who had family support and students whose fathers were farmers compared to fathers who were employed [AOR=2.71 (95% CI:1.49 - 4.92), p= 0.001].

**Conclusion:** The study found high levels of food insecurity among tertiary students in Hohoe. An insight into food insecurity among this at-risk population group requires further investigation and action. There is an urgent need for research to better understand the severity and persistence of food insecurity among higher education students in Ghana.

## Introduction

The right to adequate food is a basic human right and essential to health and well-being (1,2). Food security is defined as when all people, at all times, have physical and economic access to sufficient, safe, and nutritious food that meets their dietary needs and food preferences for an active and healthy life (3). However, Food insecurity (FI) is a collective term for inadequate access to available, affordable, and safe nutritious food. FI has become a growing public health issue worldwide and a barrier to attaining Sustainable Development Goals 1 and 2; ending poverty in all its forms and hunger by 2030 (1).

The negative consequences of FI are experienced more in low-and middle-income countries, especially in sub-Saharan Africa, Southeastern and Western Asia (4). Data has shown that approximately 60% of food insecure populations live in low-income countries (4) and about 52.5.% are in Africa (5). In Ghana, the prevalence of FI is high. For instance, in 2022 almost half of households were food insecure (6) .Food security has only increased from 5 % to 11.6 % from 2009 to 2020 (7).

An increasing number of studies have drawn attention to the high rates of food insecurity experienced on college campuses, especially in high-income and low and middle income countries (8–10). In the USA an estimated average of 14.9% of college (11) students are at risk of being food insecure, about 38–48% in Australia (12) and 17% in Europe (13). Also, up to 65% of FI rates were reported among tertiary students in South Africa (14). In Nigeria, a study showed that about 80.7 % of tertiary students were food insecure (18).

FI among college and tertiary students has received too little attention (1,15,16) especially in LMICs (17). Research has shown that college students face many challenges, which make them vulnerable to food insecurity (19,20). Students tend to perform poorly in their academics when faced with problems of food insecurity. They are more likely to make poor food choices, become overweight or obese, and be physically inactive (21,22) Moreover ,extensive financial demands such as hostel fees, utility bills, price of learning materials, and rate of tuition fees contribute to exacerbate food insecurity problems among students (19,23–25). The rising cost of higher education and delays in receiving student loans make most students in higher institutions at risk of FI (26). Other factors such as financial barriers to affordable food, lack of time for shopping and cooking, lack of cooking skills, and the struggle to have access to quality food, increases vulnerability to food insecurity (27–29). Other studies have shown that financial hardship, unemployment, high cost of student accommodation, poor skills in food management, increased reliance on borrowed money, and ineligibility for food assistance programs contribute to an increased risk of food insecurity (24,25).

Currently, there is a paucity of research on food insecurity among tertiary students in Ghana. A recent study among university students in Ghana pointed to about 60% being food insecure (30). However, this study suffers from an inadequate research design where respondents were not representative of the population under consideration and were recruited via emails resulting in low response rates.

Hohoe a city located at eastern end of Ghana, can be described as a tertiary educational hub of the Volta region. The tertiary educational institutions in the city include; two Colleges of Education, a university, and a Midwifery Training College. This presents a unique setting to assess the prevalence and associated factors of food insecurity among tertiary students in Ghana. This study provides new insight into the state of food insecurity among post-secondary students in Ghana. The study is also important for providing evidence for action on the state of food insecurity among tertiary students in Ghana.

## Materials and methods

### Study design and Sampling procedure

A descriptive cross-sectional study design was employed in assessing the prevalence of food insecurity and associated factors among tertiary students. The study was conducted on the campuses of all the tertiary institutions in the Hohoe municipality of Ghana namely, Hohoe Midwifery Training College, St. Theresa’s College of Education, St. Francis College of Education, and the University of Health and Allied Sciences (Fred Newton Binka School of Public Health). The population of interest was students aged 18 years and above. However, postgraduate students and students who were staying with their parents/ guardians were excluded from the study.

A multistage sampling method was used in selecting 411 students. The survey was conducted from June to September 2021. Before the data collection, data collection instruments were pre-tested. The structured questionnaire was sectioned into two - section A contains questions on socio-demographic characteristics, while Section B contains questions on the food security status of the respondents. The food security status of respondents over the past 12 months was assessed using an adapted 10-item United States Department of Agriculture Adult Household Food Security Survey Module (USDA HFSSM)31. A maximum time of twenty-minute was used in taking data from each respondent.

The study adhered to all ethical standards regarding human participation in research and received ethical clearance from the University of Health and Allied Sciences Research Ethics Committee (UHAS-REC A.9 12051 20-21), Ho. Participants consent were sought and each participant signed a written consent form. Also, the study complied with COVID-19 prevention protocols such as physical distancing, handwashing, and wearing masks during data collection.

### Variable and measurement

#### Food security

The food security status of respondents over the past 12 months was assessed using an adapted 10-item United States Department of Agriculture Adult Household Food Security Survey Module (USDA HFSSM). Apart from this survey tool’s wide usage and applicability, it is also less burdensome to respondents (31). A respondent’s affirmative responses (“yes, often, sometimes, almost every month, and certain months but not every month”) were scored as one and the food security score was computed as the sum of individual responses, ranging from 0-10. A response of “no” or “never” was not counted. Food security scores were divided into four categories and described as follows; (score of 0) - high food security; no limitation to food accessibility, (score of 1-2) - marginal food insecurity; one or two anxieties over food sufficiency but little or no change in their dietary intake, (score of 3-5)- low food security; low or reduced food quality or desirability of diet), (score of 6-10) - very low food security; several indications of disrupted eating patterns and reduced food intake.

Moreover, the overall scores were further categorized into either food secure or food insecure. A high and marginal food security status with 0-2 scores were categorized as food secure whereas the low and very low food security status with 3-10 scores (32) were deemed to be food insecure.

#### Anthropometry

Anthropometric measurements of respondents were conducted by trained research assistants using calibrated equipment and following a set of guidelines(33) Each respondent was weighed to the nearest 0.1 kg using a Personal digital weighing scale. Height was measured to the nearest 0.1 cm using a portable stadiometer (Seca 213). The weight and height were used to calculate the respondent’s Body Mass Index (BMI). The calculation of BMI was done by dividing weight in kilograms by height in square meters (kg/m2). The Body Mass Index of individuals was classified as underweight (<18.5), normal (18.5-24.9), overweight (25.0-29.9), and obese (≥ 30).

### Statistical Analysis

Descriptive statistics was used to explore socio-demographic characteristics and the prevalence of food insecurity. A Chi-square test was used to determine the bivariate associations between food insecurity and other variables. Variables that were associated with food insecurity significantly at the bivariate level, were further assessed with multivariate logistic regression to identify the factors associated with food insecurity.

Data generated from the questionnaires was analyzed using Stata/MP version 16.0. Study results are presented in tables, graphs, and charts. Results were reported as odds ratios at 95% confidence intervals. For all statistical tests, a p-value less than 0.05 was considered significant.

## Results

### Sociodemographic Characteristics of Respondents

A total of four hundred and eleven (411) respondents participated in this study with a mean age and standard deviation of 21.8 ±2.4 (See Table 1). The majority (64.0%) of the respondents were females. Most (98.0%) respondents were not married and without children (96.1 %). Almost half of the respondents belonged to the Ewe ethnic group (49.6%). Regarding academic levels, almost half of the respondents were in the first year (42.8%). A majority (72.6%) of the respondents were residents on-campus with approximately 67% of them in shared apartments. A greater number (53.8%) received a monthly allowance of 160-300 GHȻ and almost all (94.7%) spent an amount of 5-15 GHȻ daily on food. (That is about $0.89-2.55 at the time the data was being collected). About (97.3%) had their source of allowance from a parent/guardian and a majority (83.0%) did not receive family support for their education. Most of the respondents reported that their fathers were employed in formal/white-collared jobs (43.8%). The majority (60.1%) of their mothers were working in the informal sector. Out of the 411 respondents, (67.2%) had normal weight for height and were in the normal BMI group (24.49- 29.9 kg/m2).

**Table 1:** Bivariate Association between Sociodemographic Characteristics and Food Insecurity Among Tertiary Students.

| Variables | Food Insecure<br>[60.6%<br>(n=249)] | Food secured<br>[ 39.4%<br>(n=162)] | Total (%)<br>(n=i) | Chi-square(p-value) |
| --- | --- | --- | --- | --- |
| **School of affiliation** |  |  |  | 9.96 (0.019) |
| Franco | 68.7 (114) | 31.3 (52) | 40.4 (166) |  |
| UHAS | 54.5 (61) | 45.5 (51) | 112 (27.3) |  |
| Hohoe MTC | 46.7 (21) | 53.3 (24) | 45 (11.0) |  |
| Theresco | 60.2 (53) | 39.8 (35) | 88 (21.4) |  |
| **Sex** |  |  |  | 6.72 (0.009)** |
| Male | 68.9 (102) | 31.0 (46) | 148 (36.0) |  |
| Female | 55.9 (147) | 44.1 (116) | 263 (64.0) |  |
| **Monthly stipend** |  |  |  | 4.82 (0.090) |
| Ghc15-150 | 62.9 (78) | 37.1 (46) | 124 (30.2) |  |
| Ghc160-300 | 62.9 (139) | 37.1 (82) | 221 (53.2) |  |
| Ghc301+ | 48.5 (32) | 51.5 (34) | 66 (16.1) |  |
| **Amount spent on food** |  |  |  | 1.15(0.563) |
| GHC 5-9 | 62.7 (151) | 37.3 (90) | 241 (58.6) |  |
| GHC 10-15 | 58.1 (86) | 41.9 (62) | 148 (36.0) |  |
| GHC 16 or more | 54.6 (12) | 45.5 (10) | 22 (5.4) |  |
| **Father's occupation** |  |  |  | 16.64 (0.000)** |
| Formal | 49.4 (89) | 50.6 (91) | 180 (43.8) |  |
| Informal | 69.3 (160) | 30.7 (71) | 231 (64.2) |  |
| **Mother's occupation** |  |  |  | 6.03(0.014)** |
| Formal<sup>a</sup> | 50 (49) | 50.0 (49) | 98 (23.8) |  |
| Informal<sup>b</sup> | 63.9 (200) | 36.1 (113) | 313 (76.2) |  |
| **BMI** |  |  |  | 8.08(0.044)** |
| Underweight | 60.0 (24) | 40.0 (16) | 9.7 (40) |  |
| Healthy | 64.9 (179) | 35.1 (97) | 67.2 (276) |  |
| Overweight | 49.3 (34) | 50.7 (35) | 16.8 (69) |  |
| Obese | 46.2 (12) | 53.9 (14) | 6.3 (26) |  |
| **Family support** |  |  |  | 11.50(0.003)** |
| Yes | 56.9 (194) | 43.1 (147) | 83.0 (341) |  |
| No | 79.6 (39) | 20.4 (10) | 11.9 (49) |  |
| May be | 76.2 (16) | 23.8 (5) | 5.1 (21) |  |
| **Source of support** |  |  |  | 0.04(0.834) |
| Parents/guardian | 60.5 (242) | 39.5 (158) | 97.3 (400) |  |
| Spouse | 63.6 (7) | 36.4 (4) | 2.7 (11) |  |
| **Type of residence** |  |  |  | 3.93(0.047)** |
| On-campus | 63.3 (198) | 36.7 (115) | 76.2 (313) |  |
| Off-campus | 52.0 (51) | 47.9 (47) | 23.8 (98) |  |
| **Living arrangement** |  |  |  | 5.72(0.057) |
| Alone | 74.2 (46) | 25.8 (16) | 15.1 (62) |  |
| Roommates | 57.8 (159) | 42.2 (116) | 66.9 (275) |  |
| Parent/spouse | 59.5 (44) | 40.5 (30) | 18.0 (74) |  |
| **Mean Age in years (standard deviation)** |  |  |  | 22 ( ± 2.4) |

Abbreviation: GH¢ - Ghana Cedis, \$1= GH¢10.0, \*\*. Significant at $P \leq 0.05$

### Food Security Evaluation

Fig 1 shows the levels of food security among tertiary students. The overall prevalence of food insecurity among respondents was 60.6% and of these, 40.4% had very low food security status, 20.2 % had low food security status, 25.5% had marginal food insecurity, and 13.9 % reported high food security status over 12 months.

**Fig 1:**
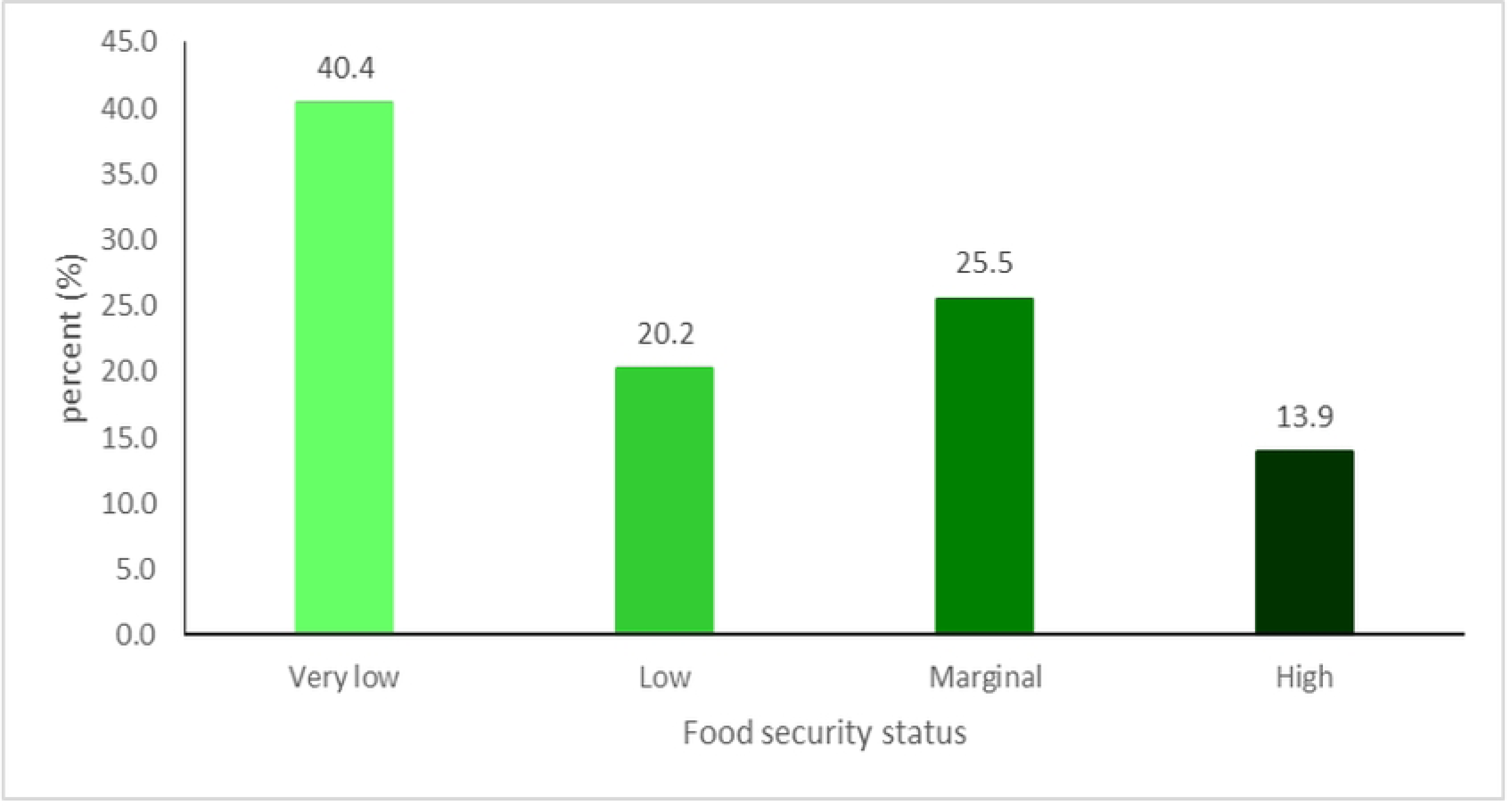
Levels of Food Security among Tertiary Students

### Bivariate Association between Sociodemographic Characteristics and Food Insecurity Among Tertiary Students

Table 1 shows results from the bivariate association between sociodemographic Characteristics and food insecurity among tertiary students. Food insecurity status was significantly associated with gender (p < 0.009), students’ residence (p < 0.047), family support (p < 0.001), father’s occupation (p < 0.001), mother’s occupation (p < 0.013), and body mass index (p <0.044)

### Multivariate analysis

In Table 2, logistic regression results of the predictors of food insecurity are presented. At the multivariate level, (Table 2) Family support was a significant factor associated with food security. Respondents who did not have support from their external family were 2 times more likely to have food insecurity as compared to those with family support. [AOR=2.56 (95% CI: 1.17 - 5.63), p= 0.019]. Also, respondents whose fathers were farmers were 2 times more likely to have food insecurity as compared to fathers who were in other forms of employment. [AOR=2.71 (95% CI: 1.49 - 4.92), p= 0.001]. Respondents whose mothers are farmers were (72%) less likely to have food insecurity than those employed but this was not statistically significant. [AOR= 1.28 (95% CI: 0.53 - 3.06), p= 0.586]. Respondents whose mothers are farmers were (72%) less likely to have food insecurity than those employed but this was not statistically significant. [AOR= 1.28 (95% CI: 0.53 - 3.06), p= 0.586]. Also, the occupations of mothers as traders and artisans were 1.19 times more likely and (52%) less likely to have food insecurity as compared to those employed. [AOR= 1.19 (95% CI: 0.70 - 2.03), p= 0.515] and [AOR= 0.48 (95% CI: 0.10 - 2.35), p= 0.365] respectively.

**Table 2:**
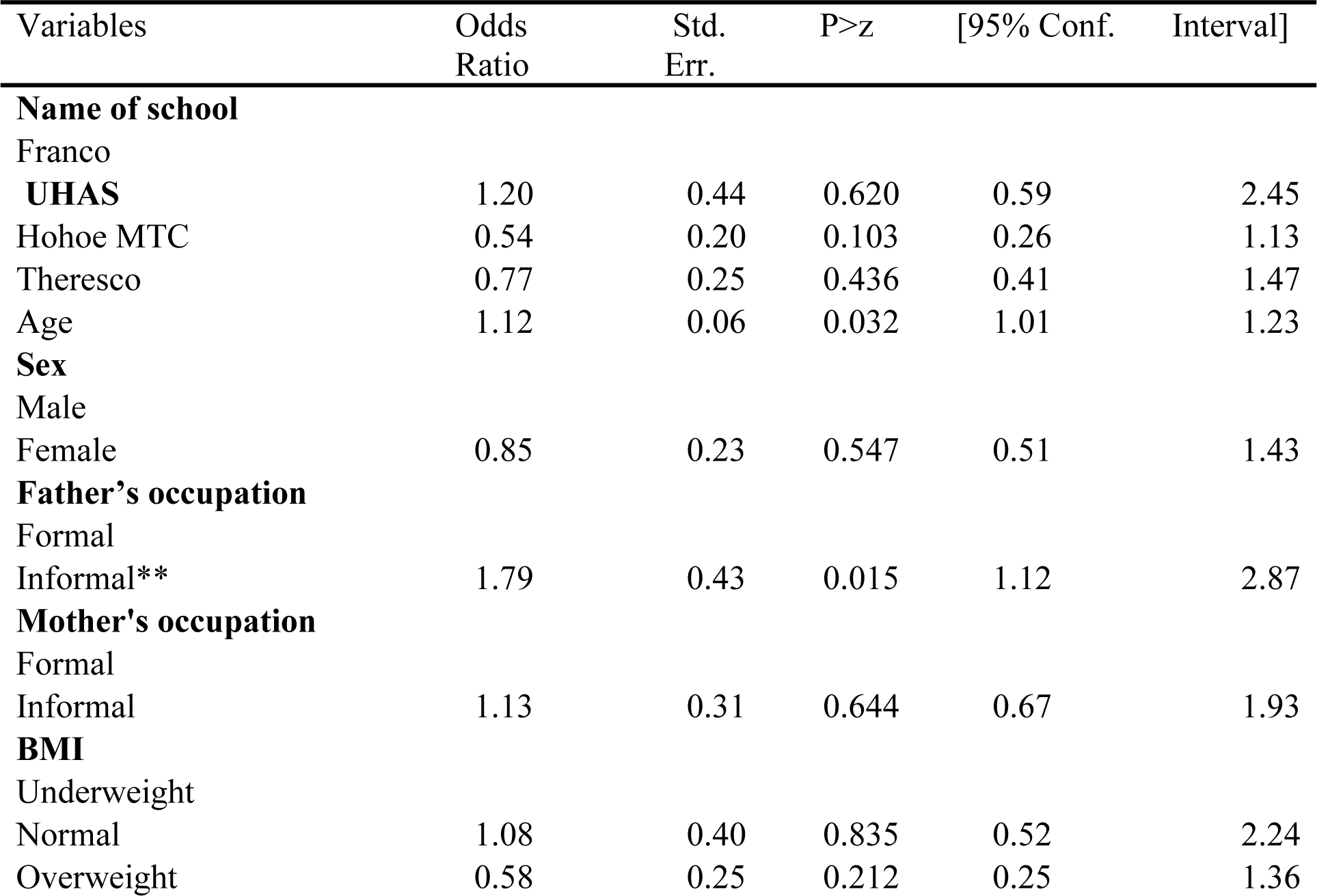
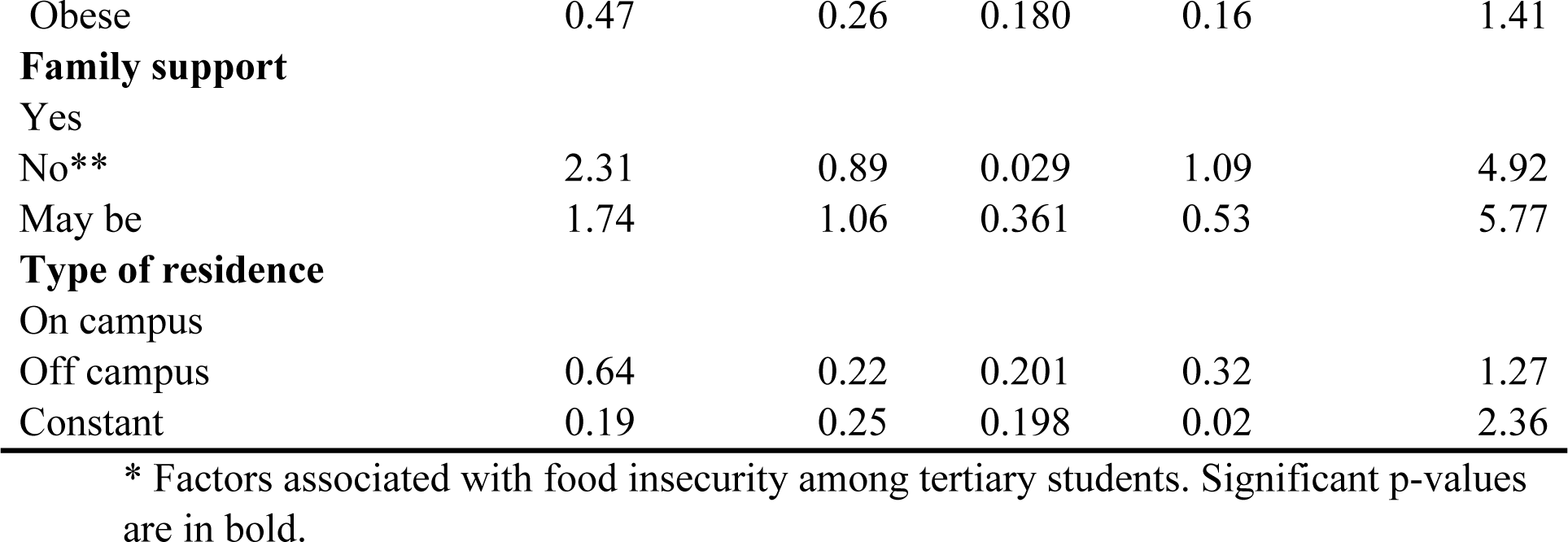
Multivariate Logistic Regression of Factors Associated with Food Insecurity Among Tertiary Students.

| Variables | Odds Ratio | Std. Err. | P>z | [95% Conf. Interval] |
| --- | --- | --- | --- | --- |
| **Name of school** |  |  |  |  |
| Franco |  |  |  |  |
| UHAS | 1.20 | 0.44 | 0.620 | 0.59 2.45 |
| Hohoe MTC | 0.54 | 0.20 | 0.103 | 0.26 1.13 |
| Theresco | 0.77 | 0.25 | 0.436 | 0.41 1.47 |
| Age | 1.12 | 0.06 | 0.032 | 1.01 1.23 |
| **Sex** |  |  |  |  |
| Male |  |  |  |  |
| Female | 0.85 | 0.23 | 0.547 | 0.51 1.43 |
| **Father's occupation** |  |  |  |  |
| Formal |  |  |  |  |
| Informal** | 1.79 | 0.43 | 0.015 | 1.12 2.87 |
| **Mother's occupation** |  |  |  |  |
| Formal |  |  |  |  |
| Informal | 1.13 | 0.31 | 0.644 | 0.67 1.93 |
| **BMI** |  |  |  |  |
| Underweight |  |  |  |  |
| Normal | 1.08 | 0.40 | 0.835 | 0.52 2.24 |
| Overweight | 0.58 | 0.25 | 0.212 | 0.25 1.36 |

|  |  |  |  |  |  |
| --- | --- | --- | --- | --- | --- |
| Obese | 0.47 | 0.26 | 0.180 | 0.16 | 1.41 |
| **Family support** |  |  |  |  |  |
| Yes |  |  |  |  |  |
| No** | 2.31 | 0.89 | 0.029 | 1.09 | 4.92 |
| May be | 1.74 | 1.06 | 0.361 | 0.53 | 5.77 |
| **Type of residence** |  |  |  |  |  |
| On campus |  |  |  |  |  |
| Off campus | 0.64 | 0.22 | 0.201 | 0.32 | 1.27 |
| Constant | 0.19 | 0.25 | 0.198 | 0.02 | 2.36 |

\* Factors associated with food insecurity among tertiary students. Significant p-values are in bold.

## Discussion

In this present study, the overall prevalence of food insecurity was found to be (60.6%) and the result suggests that food insecurity may be a common and severe issue among tertiary students in Ghana (30). This figure is higher than the national average of food insecure households (6). This finding supports evidence of food insecurity is higher among college students than households making it a public health priority. For instance, the average FI among college students stands at 43.5 % higher than 17% among households (8). FI rate ranges between 38-34 % among college students whereas household rates were around 5-10% (12,34) and a study in Portugal indicated 17. 5% among university students and 10.1 % among households (13).

In some colleges in South Africa for example, prevalence rates of 65% and 65.3% were reported among students in Free State and Kwazulu Natal Universities respectively(14,35). Further, a study among university students in Nigeria had an FI rate of 80.7%29 (18) . It is worth noting that this study was conducted immediately after the COVID-19 pandemic lockdown which worsened the food security status of most college students (32) the study showed a lower FI rate as compared to studies on the continent. A possible explanation of variability in the prevalence rate might be due to differences in the data collection instrument used in these studies (15). Also, research suggests that measuring an elusive concept like food insecurity can be difficult (36).

The current study found that the father’s occupation type was a significant factor associated with food insecurity. Students whose fathers worked in the informal sector were more likely to experience food insecurity as compared to those whose fathers were in formal employment. In Ghana, the formal sector is mostly government employees working across all sectors of the economy, and informal sector workers mostly include farmers, artisans, and traders is the largest employer. Also, students who do not receive family support showed higher odds of being food insecure. Parental occupation, especially fathers’ occupation reflects the financial capability of most students to acquire or access food since traditionally fathers are the main breadwinner of the family. In Sub-Saharan Africa, informal workers like farmers especially smallholder farmers remain among the poorest people (Castaneda et al., 2016) and it is not surprising their students whose parents work in the formal sector were likely to be food insecure. These results are consistent with a study in Nigeria where food insecurity was significantly less in students who received financial support from their parents or guardian (18). Also, another study found that students who depend only on parents or guardians for money are at an increased risk of food insecurity as compared to those with other sources of income (14).

Limited financial resources and poverty are known key predictors of food insecurity among college students (9,15,24). Several reports have also shown that financial-related factors, like student loans, low-income status, or spending habits make students more prone to food insecurity (24). Similarly, Hughes and colleagues reported that students who did not have familial financial support had increased odds of experiencing food insecurity as compared to those receiving support (38). Therefore, food vouchers, food scholarship, and cash assistants are predominant interventions used to reduce FI among college students (15).

Within the institutions of higher education assessed for food insecurity a student in the University of Health and Allied Sciences (UHAS) had higher odds of being food insecure as compared to a student at St. Francis College of Education. However, the risk of food insecurity was lower among students from Hohoe Midwifery Training College (MTC) and those from St. Theresa College of Education (Theresco). Although the associations were not statistically significant, these results give credence to how financial aid and food scholarships tend to lower FI rates. Students from the other colleges receive monthly stipends and a hot meal a day funded by the government. Although University students also have access to students’ loan, they are meager and might not be sufficient to meet the demands of university education. Studies have shown a lower risk of food insecurity among college students receiving a monthly allowance and daily expenditure(18,35,39). Another possible explanation for observed higher FI among university students than other tertiary students is the issue of food literacy. Food literacy is the term used for the practical ability of an individual able to plan, manage, select, prepare, and eat healthy foods (40). Studies have shown that FI among students of higher education is also linked to poor food literacy (15). As compared to other tertiary students in Hohoe, UHAS students do not receive hot meals and lack adequate eating places on campus.

There are various limitations to this study. The study relied on a cross-sectional study design and self-reported data from students thus subject to recall bias and misinterpretations. The study area consisted of one all-female school which may account for the overrepresentation of female students in the sample. Notwithstanding these limitations, this study makes significant contributions to food insecurity research of these at-risk population groups, given the paucity of studies among tertiary students in Ghana.

## Conclusions

The findings of this study suggest that food insecurity among tertiary students in Hohoe was high. Moreover, the results of this research showed that limited familial and financial support were the main factors associated with food insecurity. Insight into food insecurity among this at-risk population requires further investigation and action. There is a need for research to better understand the severity and persistence of food insecurity among higher education students in Ghana. Given the current situation, there is an urgent need to address food insecurity among tertiary students in Hohoe. Priority should be given to improving access, availability, and affordability of food on campus.

## Data Availability

Data for this study readily available upon request .

## Acknowledgments

We appreciate all of the participants who made this research possible.

## Notes

### Competing Interest Statement

The authors have declared no competing interest.

### Funding Statement

There was no formal funding for this project.

### Author Declarations

University of Health and Allied Sciences Research Ethics Committee

